# The practice of breast self-examination and associated factors among female healthcare professionals working in selected hospitals in Kigali, Rwanda: A Cross Section Study

**DOI:** 10.1101/2023.04.10.23288382

**Authors:** Mulugeta Tenna Wolde, Rosemary Okova, Michael Habtu, Mekitie Wondafrash, Abebe Bekele

## Abstract

**Background:** Breast self-examination is considered one of the main screening methods in detecting earlier stages of breast cancer. It is a useful technique if practiced every month by women above 20 years since globally breast cancer among women contributed to 685,000 deaths in 2020. However, the practice of breast self-examination among healthcare professionals is low in many developing countries. Therefore, this research was intended to measure the level of breast self-examination practice and determine associated factors among female healthcare professionals working in selected hospitals in Kigali, Rwanda.

**Methods:** A cross-sectional study was conducted among 221 randomly selected female healthcare professionals in four district hospitals in Kigali, Rwanda. A self-administered structured questionnaire was used as data collection instrument. The predictor variables were socio-demographic and obstetrics variables, knowledge on breast cancer and breast self-examination, attitude towards breast cancer and breast self-examination. Sample statistics such as frequencies, proportions and mean were used to recapitulate the findings in univariate analysis. Multiple logistic regression analysis was employed to identify statistically significant variables that predict breast self-examination practice. Adjusted odds ratio and 95% confidence level were reported. P-value < 0.05 was used to declare statistical significance.

**Results:** Breast self-examination was practiced by 43.5% of female healthcare professionals. This prevalence is low compared to other studies. Attitude towards breast self-examination and breast cancer was the only predictor variable that was significantly associated with breast self-examination practice [AOR=1.032; 95% CI (1.001, 1.065), p-value=0.042]. However, number of pregnancy and number of children were not significantly associated with BSE practice in the multi-variate analysis. In addition, knowledge and attitude were linearly correlated with r=0.186, p=0.005.

**Conclusions:** The breast self-examination practice was found to be low. Attitude toward breast cancer and breast self-examination was positively associated with BSE practice. Moreover, attitude and knowledge were linearly associated. This suggests the need for continuous medical education on breast self-examination and breast cancer to increase the knowledge & BSE practice level of female healthcare professionals.

## Introduction

One of the ways of improving survival rates of women with BC is early detection through screening programs (1). The three commonly practiced approaches of BC screening are clinical breast examination (CBE), mammography and breast self-examination (BSE). BSE is the most practical and useful screening method in resource scarce Sub-Saharan Africa (SSA) countries(2) where other screening methods are not widely available and practiced (3–5).

Breast self-examination is a simple, less costly physical examination done by a woman on a monthly basis privately to detect changes in texture, color, and size of her breast and also identify swelling and lumps so that earlier stages of the disease are referred to the hospital for treatment (6). The American Cancer Society (ACS) recommends that women to be cognizant of their breast look and feel and should report to a healthcare provider right away whenever they detect any abnormality (7).

According to the International Agency for Research on Cancer (IARC), in 2020 breast cancer (BC) incidence among women was estimated to be 2.3 million and contributed to 685,000 women deaths (8).

According to the study by Johns Hopkins Medical center, forty percent of breast cancers are diagnosed after females detecting a lump during routine BSE (9). Although there is still a controversy about the effectiveness of BSE in reducing cancer related mortality, WHO recommends to combine BSE with other screening methods in detecting BC at earlier stage (1).

It is reported that women in low and middle income countries don’t practice BSE widely (10). Moreover, it is negligible percentage of women who have practiced BSE with the correct procedure and timing in most African countries (11).

A study done by Ndikubwimana et al. in Kigali has shown low level of knowledge and BSE practice with correct frequency and technique (<10%) among secondary school girls in Kigali city (12). The prevalence of BSE practice among clients of health centers was 28% according to a study done in Kayonza district of Rwanda (13). The level of practice is much lower than a study done in Saudi Arabia among healthcare professionals practicing BSE, 74.7% (14). A meta-analysis done in Ethiopia has revealed 56.31% of female healthcare professionals practiced BSE(15). Similar results were also found among healthcare professionals in another study done in Ethiopia, 53% (16). This shows that the prevalence of BSE practice is higher among healthcare professionals as they are more exposed to cases of breast cancer and health education programs than the general public.

From the literatures reviewed, it can be revealed that knowledge, attitude, family history of BC and some socio-demographic characteristics (level of education, socio-economic status, and age) were regarded as good predicators for BSE practice. However, the association is not consistent as shown in the different studies. For example, the predicted variables for practicing BSE were having high level of knowledge, good attitude and having breast cancer in the family as evidenced in a study done in Africa, (4). Another predicators were personal history of BC and getting to know someone with BC (16).

A retrospective study done in Rwanda has shown that there is a delay in diagnosing breast cancer patients and most cases present in advanced stage at the start of treatment (17). Moreover, there is only one center of excellence of Butaro Hospital located in Burera District in Northern Province for treatment of all cancer patients. It is only recently that radiotherapy machines are installed in Rwanda Military Hospital (RMH) (18).

There is no adequate literature that shows the prevalence of BSE practice in Rwanda especially among healthcare professionals. Thus, the purpose of this study was to assess the practice of BSE and factors associated with BSE practice among female healthcare professionals working in selected hospitals located in Kigali, Rwanda.

## Materials and Methods

### Study Design

An institution based cross-sectional study design was employed to collect data from four district hospitals in Kigali, the capital city of Rwanda from October to December 2022. The hospitals are located in the three administrative districts of Kigali namely Gasabo (Kacyiru and Kibagabaga hospitals), Nyarugenge (Muhima hospital) and Kicukiro (Masaka hospital).

### Study Population

All female healthcare professionals aged above 20 years were eligible to be enrolled in this study. However, female healthcare professionals who had undergone a bilateral mastectomy procedure and those who declined to consent were excluded from this study.

### Sample size & sampling techniques

There were a total of 577 female healthcare workers in the four district hospitals involved in the study. A simple random probability sampling technique was utilized to select 242 samples, of which 221 female healthcare workers have returned the questionnaires. A single population proportion formula was used with assumption of a 5% significance level, a 5% precision and prevalence of 56.31% taken from a study conducted in Ethiopia (15). The 10% non-response rate and a finite population correction factor were considered to reach the final sample size of 242.

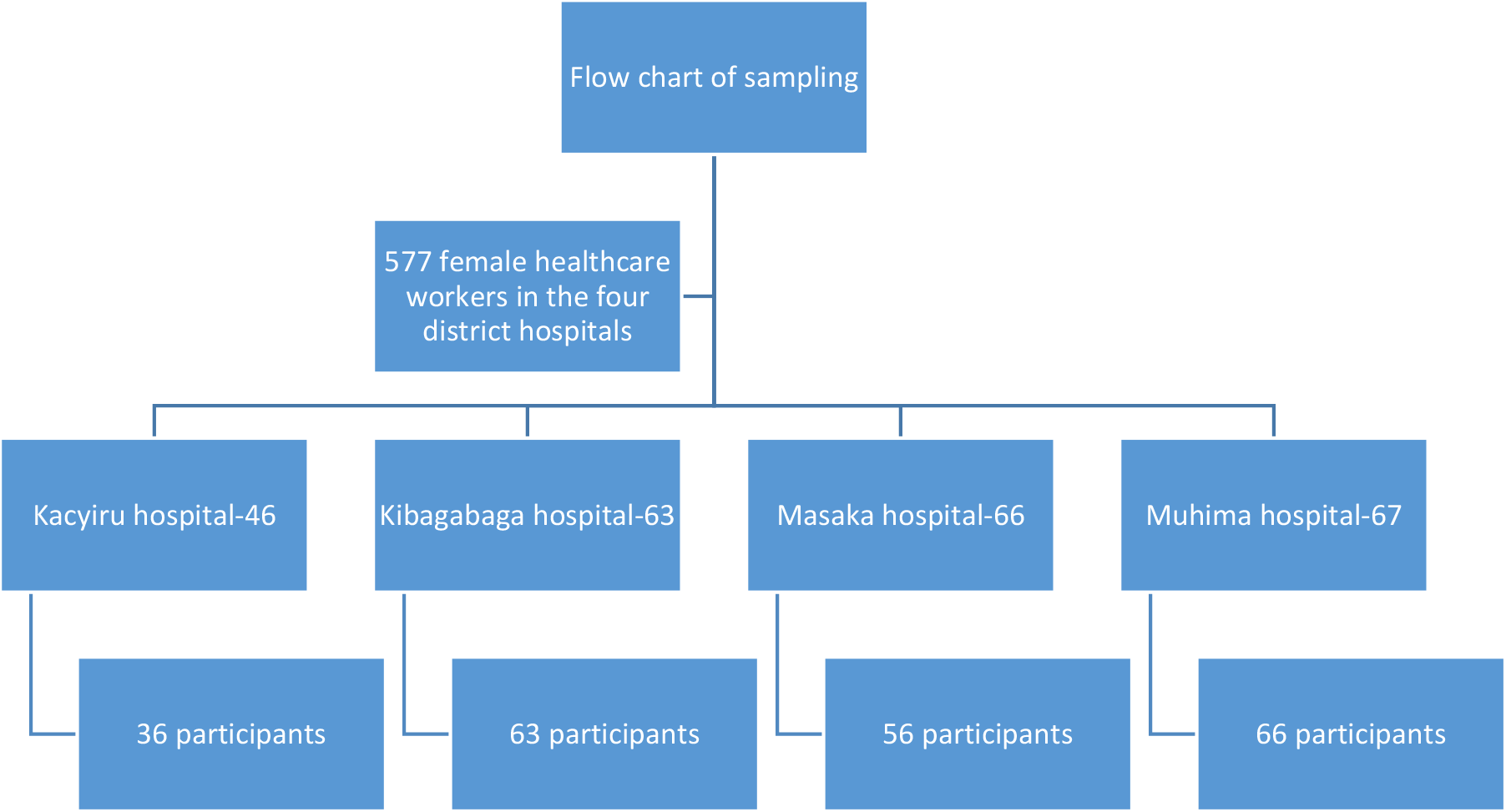

### Data quality & collection procedure

Reliability of the data collection instrument was ensured as the Cronbach’s alpha coefficient for the variable attitude was 0.89 in the pre-test and 0.8 in the actual data (19).

The data collection instrument was a pre-tested self-administered structured questionnaire. It was adapted following a detailed review of the literatures and modified to fit to this study (14). It was administered in English as most healthcare professionals in Rwanda were able to understand the language. Data collected included socio-demographic features, gynecological/obstetrics history, knowledge on BC and BSE, attitudes towards BSE and BC, practice of BSE. The only outcome variable was BSE practice while all the other variables were considered as predictor variables.

### Operational definitions

#### Attitudes

perception of the female healthcare professionals towards BSE & BC as measured by 19 elements of attitude with five points Likert scale. High score indicates a favorable attitude.

#### Good BSE Practice

a score equal to or above 7 out of 13 “Yes” or “No” questions.

#### Knowledge on BC & BSE

was defined as awareness of female healthcare professionals on BSE & BC as measured by structured knowledge questions. A high score indicates a higher knowledge.

### Data management & analysis technique

The collected data was checked for completeness and each questionnaire was coded. The paper based data was entered into Epidata v4.6 and then exported into IBM SPSS v.26 for data management, cleaning, and analysis. The total score for knowledge, attitudes and practice was calculated for each respondent where by the variables knowledge and attitude were considered as continuous variables while the construct BSE practice was considered as a categorical variable after dichotomization into Good practice and Poor practice.

Knowledge was assessed in four dimensions of BSE awareness, knowledge on BC risk factors, signs and symptoms of BC and methods of diagnosis. Each item was evaluated as ‘Yes’, ‘No’ and ‘I don’t know’. Score one (1) was given to the correct answers and zero (0) for incorrect and “I don’t know” answers (14,20). The dimension of awareness on BSE had 7 items with total score ranging from 0-7, knowledge on BC risk factors had 13 elements with total score of 0-13, BC signs and symptoms had 10 elements with a total score of 0-10 and methods of diagnosis of BC has 5 elements with a total score of 0-5. Therefore, the overall score for knowledge ranged from 0-35.

There were 19 elements for the construct attitude measured on 5-points Likert scale with a total score ranging from 0-95. Ten negatively worded items/questions in the construct attitude were reversely coded before adding the total score.

The section on BSE practice included whether the participant ever practiced BSE or not. It also included if the BSE practice was done on regular basis and as per the recommended timing and technique. The total score for BSE practice was out of 13. This variable was dichotomized as good/regular practice if the total score for BSE practice of the respondent was equal and above 7 and poor/irregular practice if the total score was below 7 (21).

Descriptive statistics such as frequencies and percentages were used to summarize the results of categorical variables while mean/standard deviation (SD) were used to summarize the findings of continuous variables in univariate analysis. Normality test was done using Shapiro-Wilk test for knowledge and attitude scores. Mann Whitney U and Kruskal Wallis tests were used to compare knowledge score as the data was not normality distributed. However, comparison for attitude was done using independent t-test and ANOVA test as the data was normally distributed.

Bivariate analysis between each independent and the BSE practice was done in binary logistic regression model and those variables that showed significant association with BSE practice with p-value of <0.25 were entered into the multivariate analysis to control for possible confounding variables and create the best fit model for prediction of the dependent variable which is practice of BSE. In multivariable binary logistic regression, adjusted odds ratio (AOR) with a 95% CI and p-values < 0.05 were considered to identify statistically significant predicators in the final model (22).

Hosmer and Lemeshow test was used to assess the goodness of fit of the data which was found to be p-value=0.73 which indicates the model adequately fitted the data. The assumption of no multi-collinearity among the continuous predictor variables was checked and the Variance Inflation Factor (VIF) was less than 5 for most of the variables that showed there was no multi-collinearity.

## Results

### Socio-demographic characteristics

A total of 242 female healthcare professionals were randomly selected and 221 returned the questionnaires with a response rate of 91.3%. The age of the respondents ranged from 22 to 58 years with mean age of 35.3 ± (SD 7.1). Close to forty-five percent of respondents 99 (44.8%) were in the age group 31-40 years of age. Of the 221 respondents, 177 (80.1%) were nurses, 10 (4.5%) were doctors and 34 (15.4%) were other medical professionals. More than half of them 127 (57.5%) were holders of advanced A1 diplomas. About 85 (38.5%) were followers of Catholic religion followed by Protestant 79 (35.7%) and Adventist 30 (13.6%). Almost three-fourth of the study population 165 (75.1%) were married. Nearly one third of the participants 77 (34.8 %) had more than 10 years of work experience. Almost all of the participants 214 (98.6%) are in the Rwandan socio-economic class (Ubudehe) category III (Table 1).

**Table 1.** Socio-demographic characteristics of female healthcare professionals in four district hospitals in Kigali Rwanda, 2022 (n=221)

| Variables | Frequency | Percent |
| --- | --- | --- |
| Age (years) |  |  |
| 21-30 | 70 | 31.7 |
| 31-40 | 99 | 44.8 |
| Above 41 | 52 | 23.5 |
| **Profession** |  |  |
| Nurse | 177 | 80.1 |
| Doctor | 10 | 4.5 |
| Other | 34 | 15.4 |
| **Level of education** |  |  |
| A2 (High-school diploma) | 11 | 5.0 |
| A1 (2-3 years college diploma) | 127 | 57.5 |
| A0 (Bachelor degree) & above | 83 | 37.6 |
| **Socio-economic category\*** |  |  |
| Category III | 218 | 98.6 |
| Category IV | 3 | 1.4 |
| **Religion** |  |  |
| Adventist | 30 | 13.6 |
| Catholic | 85 | 38.5 |
| Muslim | 6 | 2.7 |
| Protestant | 79 | 35.7 |
| Other | 21 | 9.5 |
| **Marital status** |  |  |
| Single | 48 | 21.7 |
| Married | 166 | 75.1 |
| Divorced & Widowed | 7 | 3.2 |
| **Work Experience** |  |  |
| <10 years | 144 | 65.2 |
| ≥10 years | 77 | 34.8 |

\*Category I-extremely poor, category II-poor, category III-self-sustaining, IV-rich (23)

### Obstetrics and breast cancer history of female healthcare professionals

Most of the respondents 162 (73.3%) had history of pregnancy with mean age at first pregnancy being 25.9 years ± (SD 3.5) and the average number of pregnancy and children were 2.8 ± (SD 1.5) and 2.6 ± (SD 1.2) respectively. The average duration of breastfeeding of the last child was 1.9 ± (SD 0.89) years. Minority of the participants had breast cancer history at personal level 17 (7.7%), at the levels of first degree relatives 10 (4.5%) and second degree relatives 8 (3.6%). Out of a total 54 (24.4%) participants who received training on BSE, 41.5% received on-the-job training while 35.9% at school and 22.6% in both school and on-the-job (Table 2).

**Table 2.** Obstetrics and breast cancer history of female healthcare professionals, Kigali, Rwanda, 2022, (n=221)

| **Variables** | **Mean** | **SD** | **Frequency** | **Percent** |
| --- | --- | --- | --- | --- |
| **Age at first pregnancy \*** | 25.88 | 3.50 |  |  |
| **Number of children \*** | 2.60 | 1.24 |  |  |
| **Number of pregnancy \*** | 2.83 | 1.47 |  |  |
| **Duration of breastfeeding \*** | 1.88 | .89 |  |  |
| **History of self-breast cancer** |  |  |  |  |
| No |  |  | 204 | 92.3 |
| Yes |  |  | 17 | 7.7 |
| **History of breast cancer in 1st relative** |  |  |  |  |
| No |  |  | 211 | 95.5 |
| Yes |  |  | 10 | 4.5 |
| **History of breast cancer in 2nd relative** |  |  |  |  |
| No |  |  | 213 | 96.4 |
| Yes |  |  | 8 | 3.6 |
| **Training on BSE** |  |  |  |  |
| No |  |  | 167 | 75.6 |
| Yes |  |  | 54 | 24.4 |
| **Time of BSE training received** |  |  |  |  |
| In the higher institution |  |  | 19 | 35.8 |
| On-the-job |  |  | 22 | 41.5 |
| In the higher institution and on-the-job training |  |  | 12 | 22.6 |
| **Name of the hospital** |  |  |  |  |
| Kacyiru hospital |  |  | 36 | 16.3 |
| Kibagabaga hospital |  |  | 63 | 28.5 |
| Masaka hospital |  |  | 56 | 25.3 |
| Muhima hospital |  |  | 66 | 29.9 |

\* Mean and standard deviation was calculated for continuous variables. SD=Standard Deviation

### Knowledge about breast cancer and awareness on breast self-examination

The awareness level of participants on the correct age of commencement of BSE practice was 56.6% and slightly over two-thirds of the participants 150 (67.9%) knew that BSE has to be done on monthly basis. However, only less than half of the participants 96 (43.4%) knew the recommended timing to do BSE which is 3-5 days after the cession of the monthly menses. The majority of respondents 190 (86.0%) acknowledged that BSE is used for early detection of BC and 195 (88.2%) have affirmed that it is done by inspection and palpation and 88.2% replied that a woman had to consult a doctor within few days after detecting the mass. However, the majority 177 (80.1%) wrongly reported that a healthcare professional had to wait for the progress of the mass for few months before consulting a doctor (Table 3).

**Table 3.**
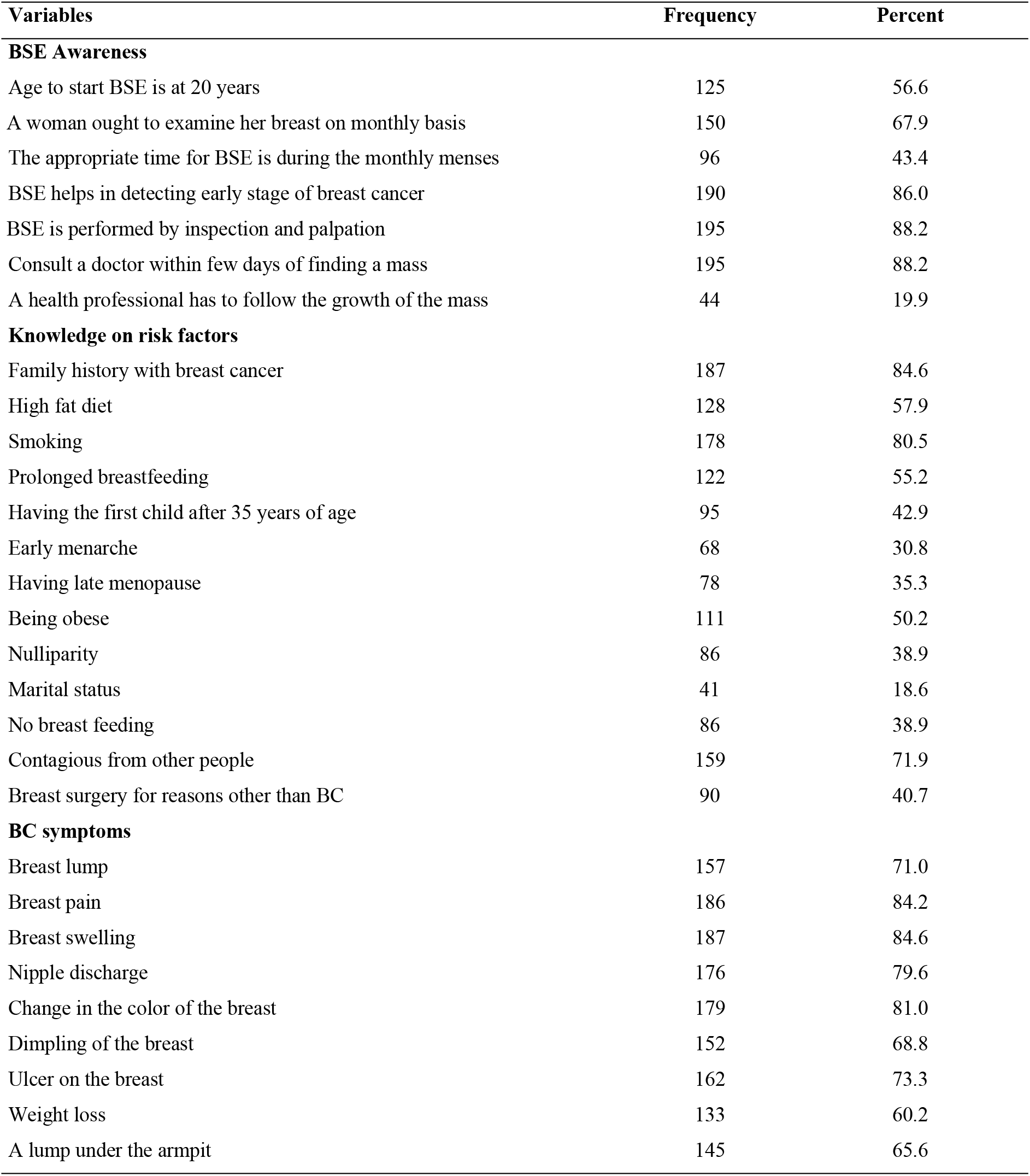
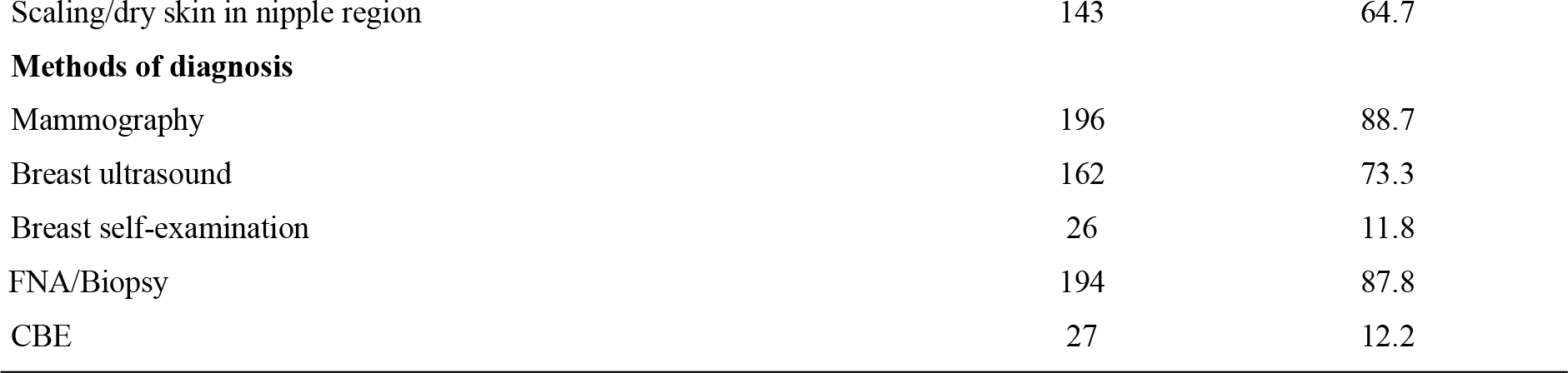
Participants correct responses on knowledge of BSE & BC questions, n=221.

The commonly recognized risk factors by female healthcare professionals were family history of breast cancer (84.6%) and smoking (80.5%). Nulliparity, having first child at older age, early menarche and late menopause were known by less than half of the participants. About 55% stated that prolonged breastfeeding is not a risk factor for BC (Table 3).

Most of the symptoms of BC were known by female healthcare professionals except weight loss that was mentioned by only 60.2% of participants. Breast pain (84.2%) and swelling (84.46%) were the most commonly recognized symptoms of breast cancer (Table 3). Most respondents knew diagnostic mammography (88.7%), breast ultrasound (73.3%) and biopsy (87.8%) as methods of diagnosis of breast cancer. However, only 12.2% and 11.8% of the respondents were aware of CBE and BSE as screening methods rather than diagnostic tools respectively (Table 3).

The overall median knowledge score of respondents was 62.9%, and only 44.8% of the study participants scored above the median. The median score of the participants on BSE awareness was 71.4% while the median score of respondents on BC risk factors, symptoms and methods of diagnosis were 46.2%, 80.0% and 60.0% respectively.

Female doctors had statistically significant higher knowledge level than nurses and other health professionals with median score of 74.3% compared to 62.9% for nurses and 58.6% for other professions, p-value=0.011 (Figure 1).

**Figure 1.**
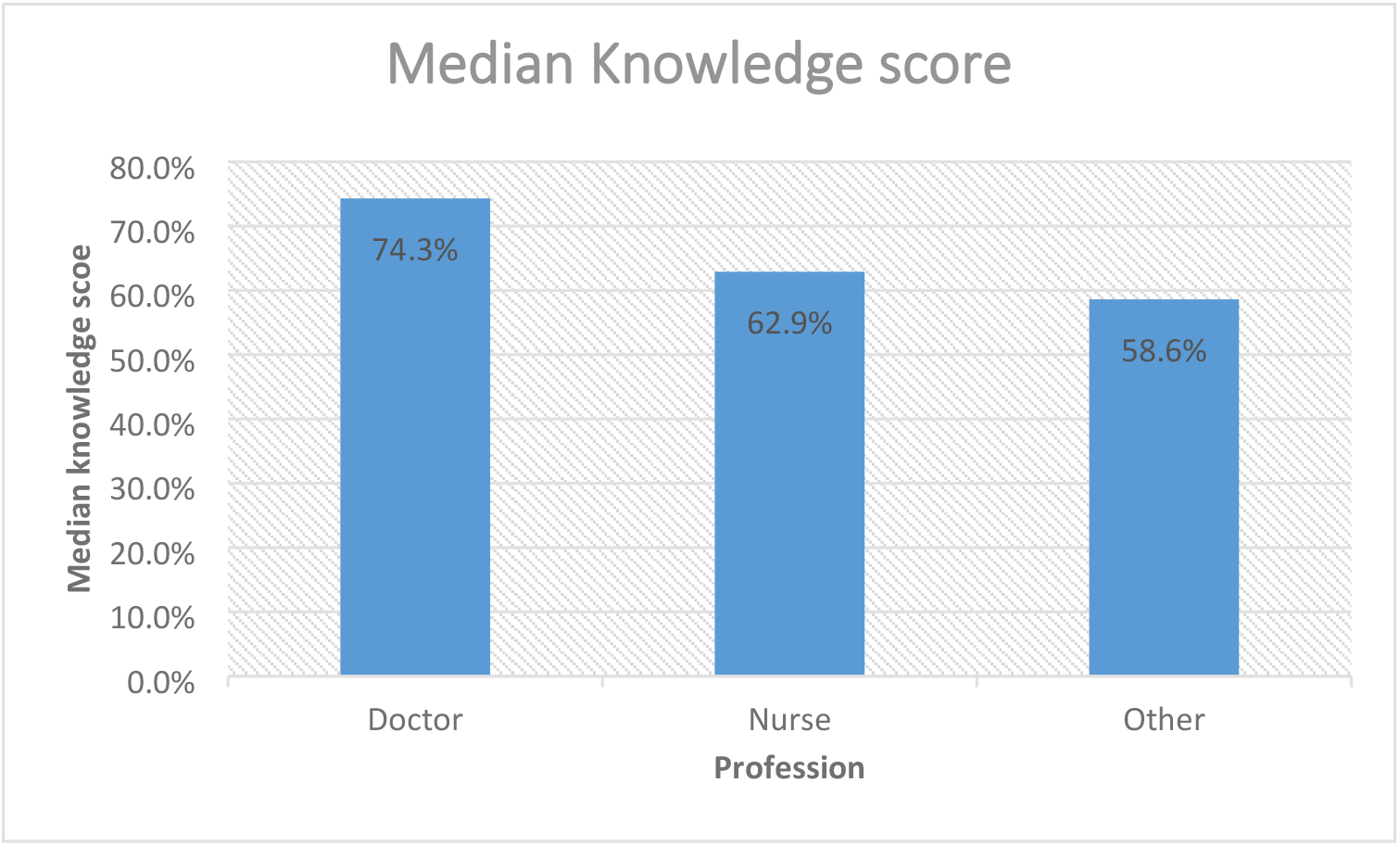
Median knowledge score among different professions, 2022

Holders of Bachelor degree (A0) had statistically significant higher total knowledge score than those with advanced diploma (A1), p=0.034.

Female healthcare professionals who received training on BSE either on the job or school had statistically significant higher total knowledge score than those without training, p-value=0.007.

There was no statistically significant knowledge difference between the groups with good BSE practice and poor BSE practice with p-value=0.064.

### Attitude of respondents to breast self-examination

The mean for the total attitude score toward BSE and BC was 72.8 ± (SD 11.1). Table 4 shows the frequency distribution of the 19 items used to measure attitude in 5-points Likert’s scale.

**Table 4.** Participants’ response to attitude towards BC and BSE, n=221.

| Attitude Questions | Likert's scale |  |  |  |  |
| --- | --- | --- | --- | --- | --- |
|  | Strongly disagree | Disagree | Neutral | Agree | Strongly agree |
| BC does not occur in the Old | 60(27.1) | 70(31.7) | 36(16.3) | 37(16.7) | 18(8.1) |
| Family history is a risk for BC | 25(11.3) | 22(10.0) | 22(10.0) | 91(41.2) | 61(27.6) |
| Advanced BC is not curable | 29(13.1) | 38(17.2) | 40(18.1) | 86(38.9) | 28(12.7) |
| BSE is a very useful method in detecting breast cancer | 21(9.5) | 13(5.9) | 21(9.5) | 101(45.7) | 65(29.4) |
| BSE should be promoted nationwide | 20(9.0) | 13(5.9) | 28(12.7) | 95(43.0) | 65(29.4) |
| I will teach women on BSE | 16(7.2) | 23(10.4) | 29(13.1) | 98(44.3) | 55(24.9) |
| Any woman can do BSE | 21(9.5) | 25(11.3) | 21(9.5) | 94(42.5) | 60(27.1) |
| There is a huge benefit from doing regular BSE | 20(9.0) | 19(8.6) | 26(11.8) | 96(43.4) | 60(27.1) |
| There are limited barriers to practice BSE | 32(14.5) | 52(23.5) | 28(12.7) | 85(38.5) | 24(10.9) |
| BSE isn't pleasant practice | 42(19.0) | 77(34.8) | 40(18.1) | 42(19.0) | 20(9.0) |
| Early diagnosis prolongs a woman's life | 16(7.2) | 24(10.9) | 25(11.3) | 97(43.9) | 59(26.7) |
| I believe that I can't identify any abnormality | 42(19.0) | 81(36.7) | 37(16.7) | 43(19.5) | 18(8.1) |
| There is no reason to do breast self-examination | 104(47.1) | 72(32.6) | 19(8.6) | 17(7.7) | 9(4.1) |
| If there is no abnormality detected during BSE, there is no need to do mammography | 50(22.6) | 100(45.2) | 21(9.5) | 35(15.8) | 15(6.8) |
| Early detection methods for breast cancer have no effect on treatment | 63(28.5) | 83(37.6) | 24(10.9) | 38(17.2) | 13(5.9) |
| BSE is very difficult for illiterate women, | 59(26.7) | 87(39.4) | 43(19.5) | 22(10.0) | 10(4.5) |
| BC is non curable | 52(23.5) | 104(47.1) | 34(15.4) | 18(8.1) | 13(5.9) |
| Only educated women can do BSE | 78(35.3) | 80(36.2) | 22(10.0) | 27(12.2) | 14(6.3) |
| BC does not occur in young | 72(32.6) | 98(44.3) | 27(12.2) | 14(6.3) | 10(4.5) |

Three quarter of the respondents 166 (74.1%) have agreed BSE as a useful method to detect breast cancer. The majority of the participants 160 (72.4%) agreed that BSE has to be promoted at the national level. Nearly two third of the respondents 154 (64.9%) believed that BSE could be done by any woman. However, only 123 (55.7%) were confident that they could identify abnormality during BSE.

Knowledge on BC and BSE had a positive linear relationship with attitude toward BSE with r=0.186, p=0.005.

### Breast self-examination practice of respondents

Less than half of the female health care professionals 94 (42.5%) have ever practiced BSE. Moreover, only 73 (33.0%) respondents have shown to have a Good/Regular practice. Only 56.4% of female healthcare professionals conducted BSE on monthly basis, and less than half of them (47.9%) practiced as per the recommended timing. Less than half of the participants (48.9%) reported to practice BSE in lying down position. However, more than 70% of participants reported to examine the arm pit, apply different pressures and follow the recommended pattern during BSE practice (Table 5).

**Table 5.** BSE Practice among healthcare professionals, n=221.

| Variables | Responses | Frequency | Percent |
| --- | --- | --- | --- |
| Ever practiced BSE | Yes | 94 | 42.5 |
| BSE done on monthly basis | Yes | 53 | 56.4 |
| BSE done few days after my monthly menses ends | Yes | 45 | 47.9 |
| BSE done standing in front of a mirror | Yes | 61 | 64.9 |
| BSE done during taking shower | Yes | 47 | 50.0 |
| BSE done in lying down position | Yes | 46 | 48.9 |
| Use the pads of three middle fingers during BSE | Yes | 63 | 67.0 |
| Examine armpit (axillary area) during breast self-examination | Yes | 74 | 78.7 |
| Follow a circular or up and down pattern during BSE | Yes | 80 | 85.1 |
| Palpate the neck region during BSE | Yes | 58 | 61.7 |
| Apply different level of pressure during BSE | Yes | 68 | 72.3 |
| Squeeze the nipple during BSE | Yes | 66 | 70.2 |
| Record the findings of BSE on a notebook | Yes | 49 | 52.1 |

Fear of being diagnosed with BC was the major reason for not practicing BSE in almost half of the participants (49.6%). A quarter of the participants (25.2%) have mentioned lack of technical knowledge as a second reason hindering BSE Practice (Figure 2).

**Figure 2.**
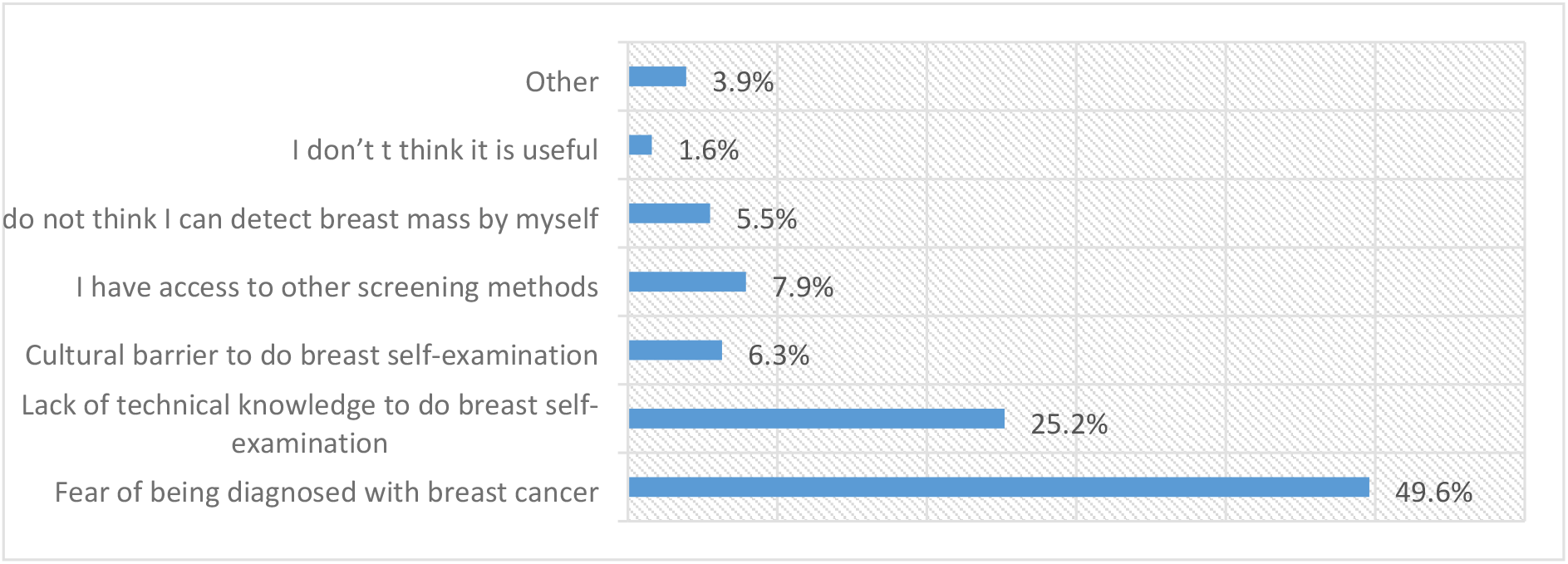
Reasons for not practicing BSE, n=127.

### Predictors of breast self-examination practice

The bivariate logistic analysis has showed that age, religion, number of pregnancy, number of children, duration of breastfeeding, training on BSE, knowledge, attitude and work experience were the candidate variables for multivariate logistic regression analysis at p-value of < 0.25.

The multivariate analysis result revealed that attitude had a significant positive impact on BSE Practice after adjusting for confounding, [AOR=1.032; 95% CI (1.001, 1.065)]. However, other candidate variables were not found to be good predictors of BSE practice (Table 6).

**Table 6.** Bivariate and Multivariate Logistic regression for association between BSE and Predictors of BSE among healthcare professionals in Kigali, Rwanda, 2022, (n=221)

| Variables | BSE Practice |  | COR(95%CI) | AOR(95%CI) | P-value. |
| --- | --- | --- | --- | --- | --- |
|  | No | Yes |  |  |  |
| **Age (years) <sup>a</sup>** |  |  | 1.023 (0.99-1.07) | 0.98 (0.91-1.06) | 0.585 |
| **Religion** |  |  |  |  |  |
| Adventist | 18 (14.2%) | 12 (12.8%) | 1 | 1 |  |
| Catholic | 39 (17.6%) | 46 (20.8%) | 1.09 (0.46-2.59) | 1.08 (0.43-2.67) | 0.287 |
| Muslim | 5 (2.3%) | 1 (0.5%) | 0.34 (0.04-3.35) | 0.33 (0.32-3.44) | 0.875 |
| Protestant | 50 (22.6%) | 29 (13.1%) | 0.80 (0.33-1.93) | 0.76 (0.30-1.94) | 0.354 |
| Other | 15 (6.8%) | 6 (2.7%) | 0.29 (0.07-1.20) | 0.29 (0.06-1.273) | 0.100 |
| **Number of pregnancy <sup>a</sup>** |  |  | 1.22 (1.04-1.43) | 1.23 (0.72-2.12) | 0.447 |
| **Number of children <sup>a</sup>** |  |  | 1.23 (1.03-1.47) | 0.91 (0.47-1.76) | 0.788 |
| **Duration of breastfeeding <sup>a</sup>** |  |  | 1.22 (0.95-1.56) | 1.03 (0.73-1.45) | 0.868 |
| **Training on BSE** |  |  |  |  |  |
| No | 100(45.2%) | 67(30.3%) | 1 | 1 |  |
| Yes | 27 (12.2%) | 27 (12.2%) | 1.74 (0.92-3.27) | 1.47 (0.74-2.91) | 0.276 |
| **Knowledge Total <sup>a</sup>** |  |  | 1.04 (0.99-1.09) | 1.02 (0.97-1.08) | 0.442 |
| **Attitude Total <sup>a</sup>** |  |  | 1.03 (1.001-1.06) | 1.03 (1.00-1.07) | 0.042 |
| **Work experience** |  |  |  |  |  |
| <10 years | 88 (39.8%) | 56 (25.3%) | 1 | 1 |  |
| ≥ 10 years | 39 (30.7%) | 38 (40.4%) | 1.53 (0.88-2.68) | 1.24 (0.46-3.32) | 0.668 |

COR=Crude Odds ratio, AOR=Adjusted Odds Ratio,
\* indicates statistical significant at p<0.05. <sup>a</sup> continuous variable. 1=Reference category

## Discussion

The present study was conducted to assess the prevalence of BSE and its associated factors among female healthcare professionals working in selected district hospitals in Kigali, Rwanda.

It was found out that the median knowledge and attitude scores of the participants were 62.9% and 73.7% respectively. Less than half (42.5%) of the female healthcare professionals practiced breast self-examination; only 33.0% showed good BSE practice. The main reasons for not practicing BSE was fear of being diagnosed with breast cancer followed by lack of technical knowledge to perform BSE. Attitude was the only statistically significant predictor for BSE practice.

Less than half of female healthcare professionals, 94 (42.5%) reported ever performing BSE, yet regular performers were only 33.0%. Similar prevalence of regular BSE practice was observed in prior study conducted among female healthcare professionals in Oromia region of Ethiopia, 32.6% (24). However, the prevalence of BSE practice in this study was lower than previous studies done in Saudi Arabia, Ethiopia, Turkey, Nigeria and Morocco (14,16,25–29).. The possible explanations for this difference could be differences in educational level of participants where most of the respondents were holders of bachelor degree in Saudi Arabia’s study. In addition, the proportion of doctors was higher in the study done by Heena in Saudi Arabia (14).

The magnitude of breast self-examination practice in this study was higher compared to a study done in North West Ethiopia (30). The study participants being young and living in relatively rural area might have an impact in the magnitude of BSE practice in Ethiopia.

Moreover, the prevalence of BSE practice in this study was higher compared to the study done among secondary students in Nyarugenge district in Kigali, Rwanda which was less than 24% and a study among women attending health facilities in Kayonza district, Rwanda 28% (12,13). This difference is mainly due to differences in the composition and educational level of the participants.

The study found out that there was no statistically significant association between the overall total knowledge score and BSE practice among female healthcare professionals. This finding was in agreement with a prior study conducted among female healthcare professionals in Nigeria (28). However, knowledge of BSE and BC was significantly associated with BSE practice in studies conducted among healthcare professionals in Ethiopia (15,24,31) and Turkey (32).

Female doctors had statistically significant higher knowledge level than nurses and other healthcare professionals with median score of 74.3% compared to 62.9% for nurses and 58.6% for other professions. This finding is supported by prior studies conducted in Morocco among healthcare professionals (29). The knowledge level of female healthcare professionals in this study was much higher than a study done in Saudi Arabia (14). This might be due to lack of updated courses and focus on BC.

Female healthcare professionals who received training on BSE on either the job or school had higher knowledge score than those without training.

Holders of Bachelor degree (A0) had statistically significant higher total knowledge score than advance diploma (A1). This is in consistent with a prior study done in Oromia region of Ethiopia (24).

Participants’ attitude towards BSE and BC was significantly associated with BSE practice. This finding was supported by prior studies done in Turkey (33). However, this is in contrary to the results obtained in studies carried out in Nigeria (29) and Morocco (29) where attitude was not associated with practice.

There was statistically significant differences in the total attitude scores among the different professions. Doctors had a higher attitude score than nurses and other professions with median scores of 86.8%, 72.6%, and 73.7% respectively. However, the attitude score didn’t differ significantly among the different level of education.

The multivariate analysis has shown that attitude towards BSE and BC as the only significant predictor variable to perform BSE. This finding was supported by prior studies done in Ethiopia (15).

In addition, knowledge and attitude had a positive linear relationship with r=0.186, p=0.005.

There were certain limitations in the study. The first was the practice of BSE was assessed by self-reporting by the respondents. This might not provide the actual facts as some respondents might not adequately remember the timing and frequency of practice. The second limitation was that respondents were female healthcare professionals working in the district hospitals, this result may not reflect those working in the health centers and private institutions. Despite this limitation, the study identified important gaps in knowledge and practice of BSE among healthcare professionals in Kigali, Rwanda.

## Conclusions

The median knowledge and attitude scores of the participants were 62.9% and 73.7% respectively. Moreover, only less than half (42.53%) of the female healthcare professionals practiced BSE. The main reason for not practicing BSE was fear of being diagnosed with breast cancer. Attitude was the only predictor for BSE practice that was statistically significant.

The study recommends the Ministry of Health and Rwanda Biomedical Center to organize trainings for healthcare professionals who are considered as role models for the public to fill the knowledge gap and promote early detection of breast cancer among the health professionals and in the society at large. The study suggests for further research to be carried out that involves both quantitative and qualitative approaches in order to understand the causes for low knowledge and prevalence of BSE practice among healthcare professionals.

## Data Availability

The dataset used and analyzed for the current study is not publicly available because we are planning to produce other papers. However it is available from the corresponding author on reasonable request.

## Acronyms and abbreviations

ACS: American Cancer Society
BC: Breast Cancer
BSE: Breast Self-Examination
CBE: Clinical Breast Examination
CI: Confidence Interval
IARC: International Agency for Research on Cancer
RMH: Rwanda Military Hospital
SPSS: Statistical Package for Social Sciences
SSA: Sub-Saharan Africa
WHO: World Health Organization

## Acknowledgements

The authors acknowledge the interest and participation of the female healthcare professionals, and the authorities of the sampled district hospitals, for permitting to carry out the study.

## Author Contributions

**Conceptualization**: Mulugeta Tenna Wolde, Rosemary Okova, Michael Habtu.

**Data curation**: Mulugeta Tenna Wolde.

**Formal analysis**: Mulugeta Tenna Wolde

**Funding acquisition**: NA.

**Investigation**: Mulugeta Tenna Wolde.

**Methodology**: Mulugeta Tenna Wolde, Rosemary Okova, Michael Habtu.

**Project administration**: Mulugeta Tenna Wolde, Rosemary Okova, Michael Habtu.

**Resources**: Mulugeta Tenna Wolde.

**Software**: Mulugeta Tenna Wolde.

**Supervision**: Rosemary Okova, Michael Habtu, Abebe Bekele, Mekitie Wondafrash.

**Validation**: Abebe Bekele, Mekitie Wondafrash.

**Visualization**: Mulugeta Tenna Wolde.

**Writing – original draft**: Mulugeta Tenna Wolde.

**Writing – review & editing**: Rosemary Okova, Michael Habtu, Abebe Bekele, Mekitie Wondafrash.

## Supporting information

## References

1. WHO. Guidelines for the Early Detection and Screening of Breast Cancer. WHO World Heal Organ. 2016;12(3):1–57.

2. Fondjo LA, Owusu-Afriyie O, Sakyi SA, Wiafe AA, Amankwaa B, Acheampong E, et al. Comparative Assessment of Knowledge, Attitudes, and Practice of Breast Self-Examination among Female Secondary and Tertiary School Students in Ghana. Int J Breast Cancer. 2018;2018.

3. Semiglazov’ VF, Moiseenko2 VM. Breast self-examination for the early detection of breast cancer: a USSR/WHO controlled trial in Leningrad. Vol. 65, Bulletin of the World Health Organization. 1987.

4. Seifu W, Mekonen L. Breast self-examination practice among women in Africa: a systematic review and Meta-analysis. Arch Public Heal [Internet]. 2021 Dec 1 [cited 2021 Dec 27];79(1). Available from: https://doi.org/10.1186/s13690-021-00671-8

5. Mihret MS, Gudayu TW, Abebe AS, Tarekegn EG, Abebe SK, Abduselam MA, et al. Knowledge and Practice on Breast Self-Examination and Associated Factors among Summer Class Social Science Undergraduate Female Students in the University of Gondar, Northwest Ethiopia. J Cancer Epidemiol. 2021;2021:1.

6. Johnson O. Awareness and practice of breast self examination among women in different African countries: A 10-year review of literature. Niger Med J. 2019;60(5):219.

7. ACS. ACS Breast Cancer Screening Guidelines [Internet]. 2019 [cited 2022 Mar 15]. Available from: https://www.cancer.org/cancer/breast-cancer/screening-tests-and-early-detection/american-cancer-society-recommendations-for-the-early-detection-of-breast-cancer.html

8. Sung H, Ferlay J, Siegel RL, Laversanne M, Soerjomataram I, Jemal A, et al. Global Cancer Statistics 2020: GLOBOCAN Estimates of Incidence and Mortality Worldwide for 36 Cancers in 185 Countries. CA Cancer J Clin. 2021;71(3):209–49.

9. Cancer AB, Support F. Breast Self-Exam Detecting Breast Cancer Earlier. 2021;1–13.

10. Pengpid S, Peltzer K. Knowledge, attitude and practice of breast self-examination among female university students from 24 low, middle income and emerging economy countries. Asian Pacific J Cancer Prev. 2014;15(20):8637–40.

11. Sama CB, Dzekem B, Kehbila J, Ekabe CJ, Vofo B, Abua NL, et al. Awareness of breast cancer and breast self-examination among female undergraduate students in a higher teachers training college in Cameroon. Pan Afr Med J. 2017;28:1–9.

12. Ndikubwimana J, Nyandwi JB, Mukanyangezi MF, Kadima JN. Breast Cancer and Breast Self-examination: Awareness and Practice among Secondary School Girls in Nyarugenge District, Rwanda. Int J Trop Dis \& Heal. 2015;1–9.

13. Igiraneza PC, Omondi LA, Nikuze B, Uwayezu MG, Fitch M, Niyonsenga G. Factors influencing breast cancer screening practices among women of reproductive age in South Kayonza District, Rwanda. Can Oncol Nurs J [Internet]. 2021 Jul 22 [cited 2022 Jan 22];31(3):251. Available from: /pmc/articles/PMC8320799/

14. Heena H, Durrani S, Riaz M, Alfayyad I, Tabasim R, Parvez G, et al. Knowledge, attitudes, and practices related to breast cancer screening among female health care professionals: A cross sectional study. BMC Womens Health. 2019 Oct 22;19(1):1–11.

15. Mekonnen BD. Breast self-examination practice and associated factors among female healthcare workers in Ethiopia: A systematic review and meta-analysis. PLoS One [Internet]. 2020;15(11 November):1–18. Available from: http://dx.doi.org/10.1371/journal.pone.0241961

16. Habtegiorgis SD, Getahun DS, Telayneh AT, Birhanu MY, Feleke TM, Mingude AB, et al. Ethiopian women’s breast cancer self-examination practices and associated factors. A systematic review and meta-analysis. Cancer Epidemiol [Internet]. 2022;78(June):102128. Available from: https://doi.org/10.1016/j.canep.2022.102128

17. Uyisenga JP, Butera Y, Debit A, Josse C, Costas &, Ainhoa C, et al. Prevalence of Histological Characteristics of Breast Cancer in Rwanda in Relation to Age and Tumor Stages. Horm Cancer [Internet]. 2020 Oct 1 [cited 2021 Jun 20];11(5–6):240–9. Available from: https://doi.org/10.1007/s12672-020-00393-3

18. Rubagumya F, Costas-Chavarri A, Manirakiza A, Murenzi G, Uwinkindi F, Ntizimira C, et al. State of Cancer Control in Rwanda: Past, Present, and Future Opportunities. JCO Glob Oncol. 2020;(6):1171–7.

19. Roberts P, Priest H. Reliability and validity in research. Nurs Stand. 2006;20(44):41–5.

20. Andegiorgish AK, Kidane EA, Gebrezgi MT. Knowledge, attitude, and practice of breast Cancer among nurses in hospitals in Asmara, Eritrea. BMC Nurs. 2018;17(1):1–7.

21. Paulsamy P, Alshahrani SH, Qureshi AA, Sampayan ELE, Venkatesan K, Sethuraj P. Breast Self-examination: Knowledge, Attitude and Practice among Female College Students. J Pharm Res Int. 2021;33:460–5.

22. Shah A, Haider G, Abro N, Hashmat S, Chandio S, Shaikh A, et al. Correlation Between Site and Stage of Breast Cancer in Women. Cureus. 2022;14(2).

23. Williams TP, Nzahabwanayo S, Lavers T, Ndushabandi E. Distributing Social Transfers in Rwanda: The Case of the Vision 2020 Umurenge Programme (VUP). SSRN Electronic Journal. 2020. 1–32 p.

24. Ahmed Shallo S, Dori Boru J. Breast self-examination practice and associated factors among female healthcare workers in West Shoa Zone, Western Ethiopia 2019: a cross-sectional study. BMC Res Notes [Internet]. 2019;12:637. Available from: https://doi.org/10.1186/s13104-019-4676-3

25. Elias LN, Worku DH, Alemu SM. Assessment of breast self-examination practice and associated factors among female health professionals in Western Ethiopia: A cross sectional study. Int J Med Med Sci. 2017;9(12):148–57.

26. Akpinar YY, Baykan Z, Naçar M, Gün I, Çetinkaya F. Knowledge, attitude about breast cancer and practice of breast cancer screening among female health care professionals: A study from Turkey. Asian Pacific J Cancer Prev. 2011;12(11):3063–8.

27. Duymuş ME, Aydın HA. Original Article Evaluation of Awareness, Behavior, and Knowledge Levels of Female Healthcare Professionals About Breast and Cervical Cancer in Southern Turkey Türkiyenin Güneyindeki Kadın Sağlık Çalışanlarının Meme ve Rahim Ağzı Kanseri Konusundaki Far. 2022;87–100.

28. Ibrahim NA, Odusanya OO. Knowledge of risk factors, beliefs and practices of female healthcare professionals towards breast cancer in a tertiary institution in Lagos, Nigeria. BMC Cancer. 2009;9:1–8.

29. Ghanem S, Glaoui M, Elkhoyaali S, Mesmoudi M, Boutayeb S, Errihani H. Knowledge of risk factors, beliefs and practices of female healthcare professionals towards breast cancer, Morocco. Pan Afr Med J. 2011;10:1–10.

30. Dagne AH, Ayele AD, Assefa EM. Assessment of breast self-examination practice and associated factors among female workers in Debre Tabor Town public health facilities, North West Ethiopia, 2018: Cross-sectional study. PLoS One. 2019 Aug 1;14(8).

31. Tesfaw A, Tiruneh M, Tamire T, Yosef T. Factors associated with advanced-stage diagnosis of breast cancer in north-west Ethiopia: a cross-sectional study. Ecancermedicalscience. 2021;15.

32. Erdem Ö, Toktag E. Breast Self-Examination and Mammography among Female Primary Healthcare Workers in DiyarbakJr, Turkey. 2016; Available from: http://dx.doi.org/10.1155/2016/6490156

33. Ertem G, Koçer A. Breast self-examination among nurses and midwives in Odemis health district in Turkey. Indian J Cancer. 2009;46(3):208–13.

